# Real-time assessment of COVID-19 impact on global surgical case volumes

**DOI:** 10.1101/2020.05.03.20086819

**Authors:** Vikas N. O’Reilly-Shah, Wil Van Cleve, Dustin R. Long, Vanessa Moll, Faye M. Evans, Jacob E. Sunshine, Nicholas J. Kassebaum, Ewen M Harrison, Craig S. Jabaley

**Affiliations:** Department of Anesthesiology & Pain Medicine, University of Washington School of Medicine, 1959 NE Pacific Street, Seattle, WA 98195; Institute of Anesthesiology, University Hospital Zurich, Raemistrasse 100, 8032 Zurich, Switzerland; Department of Anesthesiology, Critical Care, and Pain Medicine, Boston Children’s Hospital, Harvard Medical School, Boston MA, USA; Dept of Health Metrics Sciences, University of Washington, Seattle, WA, USA; Dept of Global Health, University of Washington, Seattle, WA, USA; Institute for Health Metrics and Evaluation, University of Washington, Seattle, WA, USA; Centre for Medical Informatics, Usher Institute, University of Edinburgh Secretary: 0131 242 3614 (Susan Keggie, Administrative Assistant) and 0131 242 3616 (Murray Britton, Administrative Secretary); Department of Anesthesiology, Emory University School of Medicine, 1364 Clifton Road, NE, Atlanta, GA, 30322

## Abstract

**Importance:** The COVID-19 pandemic has disrupted global surgical capacity. The impact of the pandemic in low and middle income countries has the potential to worsen already strained access to surgical care. Timely assessment of surgical volumes in these countries remains challenging.

**Objective:** To determine whether usage data from a globally used anesthesiology calculator mobile application can serve as a proxy for global surgical case volume and contribute to monitoring of the impact of the COVID-19 pandemic, particularly in World Bank low income countries where official data collection is not currently practical.

**Design:** Subset of data from an ongoing observational cohort study of users of the application collected from October 1, 2018 to April 18, 2020.

**Setting:** The mobile application is available from public sources; users download and use the application per their own clinical needs on personal mobile devices.

**Participants:** No user data was excluded from the study.

**Exposure(s):** Events with impacts on surgical case volumes, including weekends, holidays, and the COVID-19 pandemic.

**Main Outcome(s) and Measure(s):** It was previously noted that application usage was decreased on weekends and during winter holidays. We subsequently hypothesized that more detailed analysis would reveal impacts of country-specific or region-specific holidays on the volume of app use.

**Results:** 4,300,975 data points from 92,878 unique users were analyzed. Physicians and other anesthesia providers comprised 85.8% of the study population. Application use was reduced on holidays and weekends and correlated with fluctuations in surgical volume. The COVID-19 pandemic was associated with substantial reductions in app use globally and regionally. There was strong cross correlation between COVID-19 case count and reductions in app use. By country, there was a median global reduction in app use to 58% of baseline (interquartile range, 46%-75%). Application use in low-income continues to decline but in high-income countries has stabilized.

**Conclusions and Relevance:** Application usage metadata provides a real-time indicator of surgical volume. This data may be used to identify impacted regions where disruptions to surgical care are disproportionate or prolonged. A dashboard for continuous visualization of these data has been deployed.

**Key Points:** *Question:* Can usage data from a globally used anesthesiology calculator mobile application contribute to monitoring of the impacts to global surgical case volume caused by the COVID-19 pandemic, particularly in resource-limited environments such as World Bank low income countries?

*Findings:* In this ongoing observational cohort study, application usage data from 92,878 unique users in 221 countries was found to serve well as a qualitative proxy for surgical case volume, with clear impacts to app use during weekend, holidays, and during the COVID-19 pandemic.

*Meaning:* This proxy of surgical volume will provide insight into the impact of and recovery from the COVID-19 pandemic where official data collection is not currently practical. A real time dashboard tracking this proxy of global surgical volume is live and under continued development.

## Introduction

The COVID-19 pandemic has resulted in substantial disruptions to healthcare delivery due to resource limitations, supply chain disruptions, the need to protect or backfill sickened healthcare workers, social distancing, and the realities of meeting surge demands. Many healthcare systems have responded by canceling or delaying elective surgical procedures.^1–4^ The downstream impacts on public health due to delays in diagnostic and therapeutic procedural care are unknown. The Lancet Commission on Global Surgery identified a profound gap in the availability of safe anesthetic and surgical care in low- and middle-income countries (LMIC), with an estimated 4.8 billion people lacking access to surgery at baseline.^5,6^ While high-income countries (HICs) are better suited to absorb disruptions in surgical care, the effect of the COVID-19 pandemic on unmet needs in LMICs could be devastating. The time course of recovery to baseline conditions following the pandemic may also be prolonged due to depletion of healthcare resources in LMICs due to their limited capacity for resource recovery.

Assessing the volume of global surgical care is notoriously difficult. Prior work in this area has relied on estimation based on modeling and labor-intensive retrospective analysis of data from nations where such information is routinely recorded and available.^5,7–9^ No real-time data sources for LMIC surgical volume are presently available.

We previously developed a free anesthesia calculator mobile application (app) for the Android platform. Called “Anesthesiologist”, it has been in use since 2011 and has garnered worldwide adoption, with over two hundred thousand users from nearly every country in the world.^10^ As previously described, the primary user base of this application are physician anesthesiologists and other anesthesia providers.^10^ The app provides practical information about airway management and drug dosing and is used for decision support in the clinical care of patients. The app has substantially greater use in LMICs as compared to HICs.^10^ Our goal in this study was to determine if utilization of the app, aggregated over the large existing international user base, could serve as a real-time qualitative proxy for surgical case volume, which could be used to monitor the impact of, and recovery from, the COVID-19 pandemic.

## Methods

### Ethics Approval and Manuscript Preparation

The study was reviewed and approved by the Emory University Institutional Review Board (study number 00082571), and there is a reliance agreement in place with the University of Washington Institutional Review Board. The approval includes a waiver of written informed consent. Participants anonymously gave electronic consent before participating in any data collection. The app is a medical device that falls into the category of enforcement discretion per the United States Food and Drug Administration.^11^ This manuscript was prepared in accordance with the STROBE checklist for improved reporting of outcomes from observational studies.^12^

### Data Collection

Data collection using the app has previously been described.^10,13^ In brief, the app provides anesthesia references and drug calculation capabilities, and following development was deployed on the Google Play Store platform.^14,15^ The app provides us with integrated data regarding service utilization and as well as responses to user surveys.^16^ The anonymized information collected includes timestamps from the mobile devices, time zone information, basic demographics, user location based on three sources (global positioning system; internet protocol address^17^; subscriber identity module country code), and app usage patterns. This data is collected and collated as previously described.^10,13^ The approach to survey data collection allowed users to opt out at any time, resulting in survey fatigue and missing data, impacting missingness for demographic data collected.^18^ Data analyzed in the present work were collected between October 1, 2018 and April 18, 2020 (final full day of data collection). The electronic data warehouse at the University of Washington and at Seattle Children’s Hospital were queried for aggregate surgical case counts which were used for the generation of panels shown in Online Supplement, Figure S2 and Figure S3.

### Statistical Methods

Raw data were downloaded and analyzed in R using RStudio v3.6.2 (R Core Team, Vienna, Austria).^19,20^ Data containing invalid timestamps were removed (280 of 4,300,975 observations). Time series data were calculated from incoming individual data points (including logged app uses and in-app navigation) from all users. World Bank classification of country income level and region was made based on publicly available classification as of April 2020.^21^ The European Centers for Disease Control was queried for data related to global COVID-19 impact (case counts, death counts).^22^ Muslim-majority countries were defined as the set of 38 countries with greater than 80% of the population characterized as observing Islam (Online Supplement, Table 1). Change point detection in time series data were performed with the cpm package using raw time series data.^23^ For clarity of presentation, plots in Figure 1 were generated using a 7 day moving average to mitigate the routine effect of app use reductions occurring on weekends (Online Supplement, Figure S1). Country-level reductions in rate of app use were generated by (a) taking the mean daily counts for April 13, 2020 through April 18, 2020; (b) taking the mean daily counts of app use from Sep 1, 2020 through Nov 1, 2020; and then calculating (a)/(b), yielding the app use for the most recent data available as a percent of baseline use. The choropleth in Online Supplement, Figure S4 was generated using the tmap package for R.^24^

**Figure 1:**
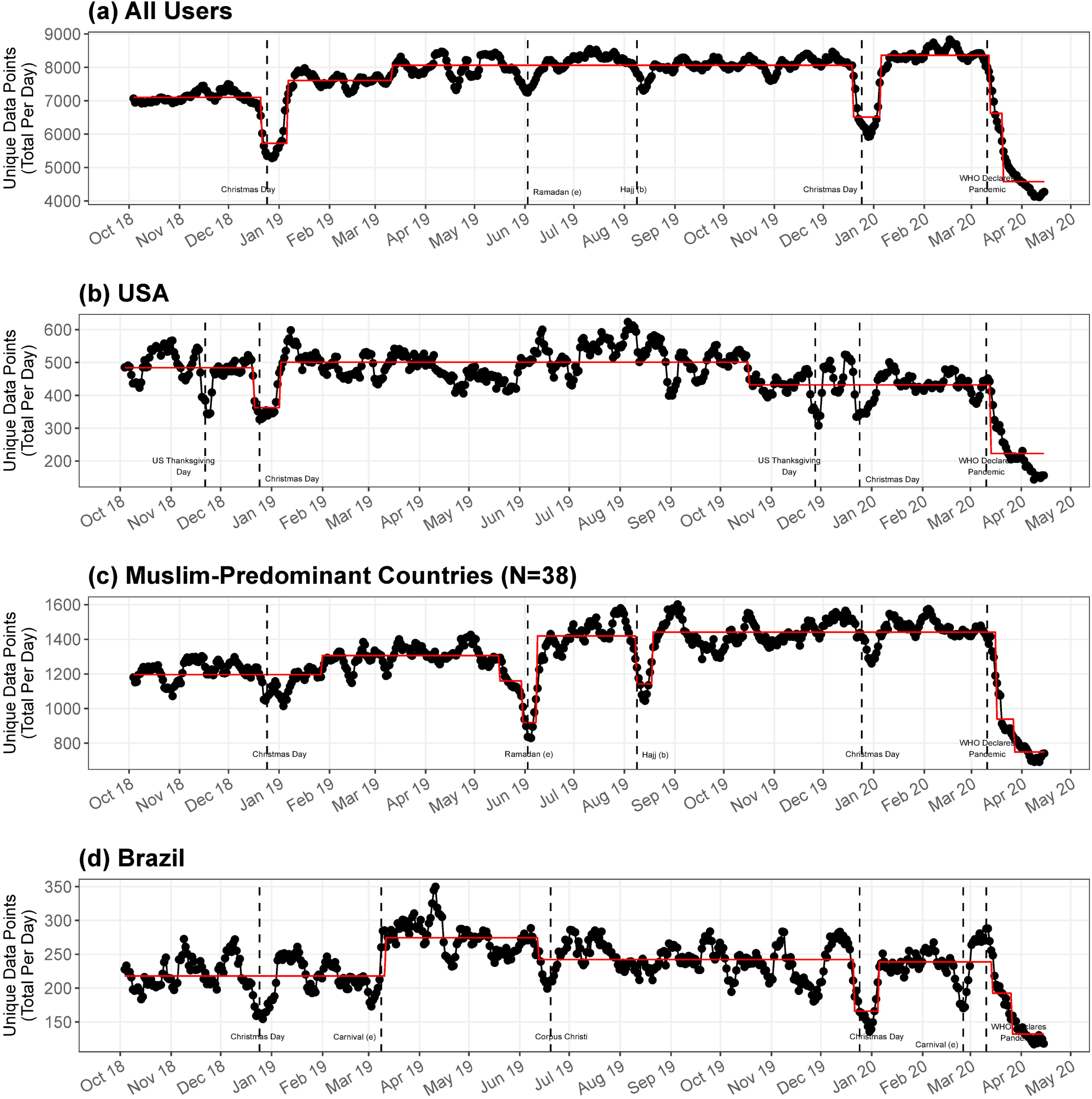
Time series data demonstrating app use for (a) all users and in specific regions: (b) USA, (c) Muslin-predominant countries, (d) Brazil. Dates of key holidays are indicated by vertical dashed lines.

## Results

### Demographics

From October 1, 2018 through April 18, 2020, there 4,300,975 data points collected and analyzed from 92,878 unique users. Provider demographics are summarized in Table 1. Incomplete survey completion resulted in partial data available from some users. The majority of users were anesthesia providers: physicians, certified registered nurse anesthetists, or anesthesiologist assistants. As discussed in a previous publication, anesthesia officers practicing in low-income countries may have identified as technically trained in anesthesia or otherwise self-identified.^10^ There was a wide distribution in terms of self-reported elements of practice environment. The distribution of participant characteristics are consistent with previous findings.^10^

**Table 1:**
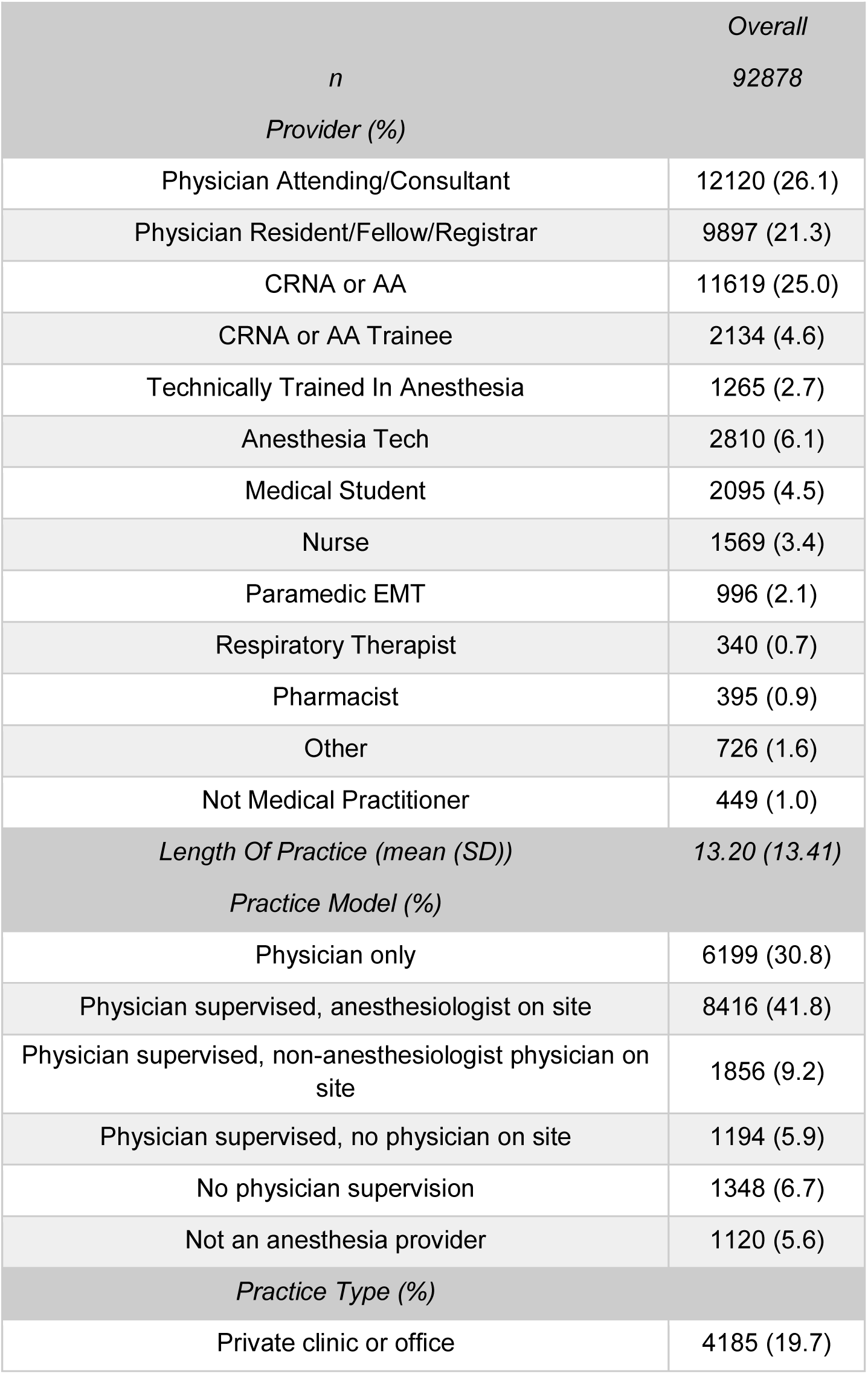
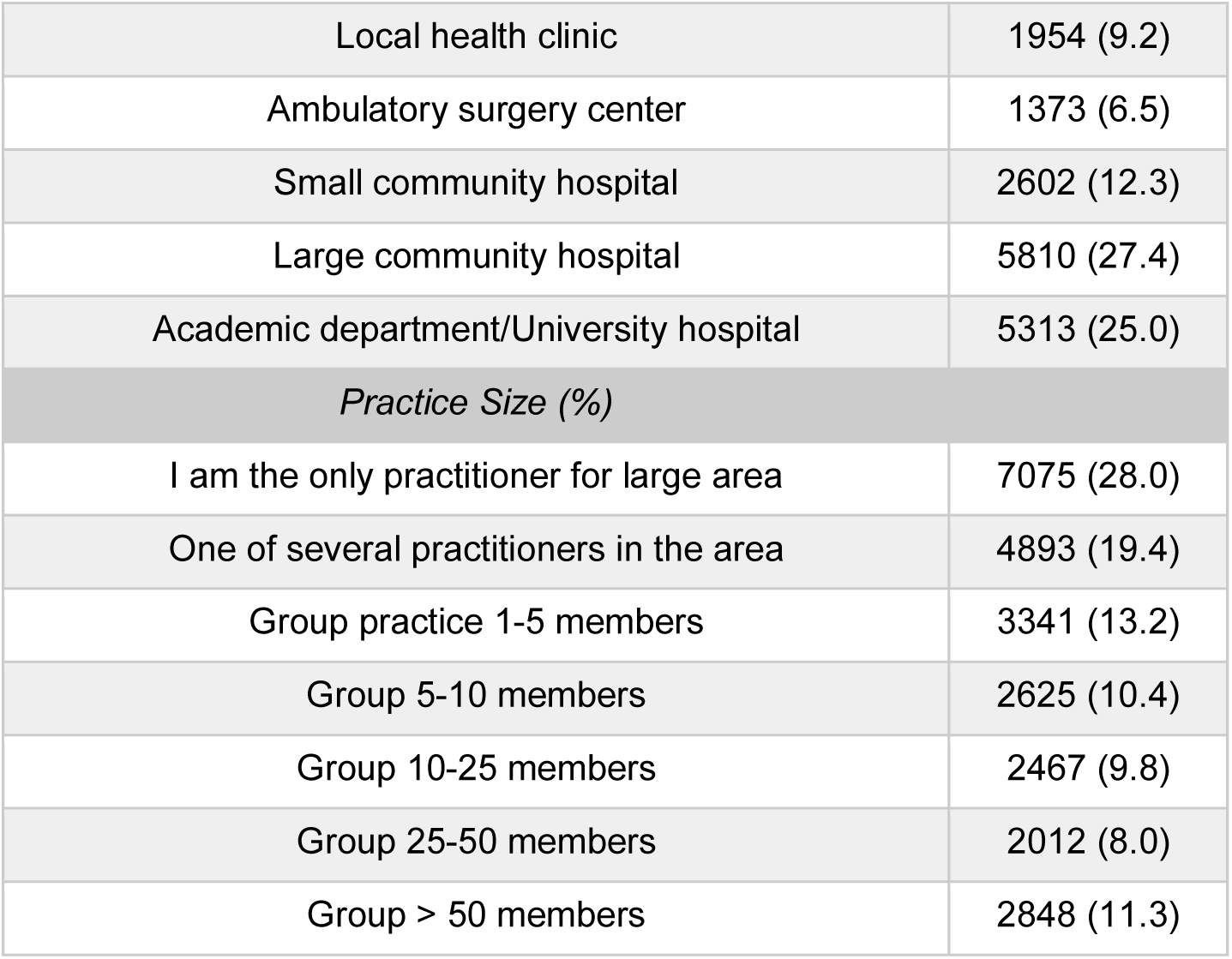
App user characteristics. CRNA=Certified Registered Nurse Anesthetist; AA=Anesthesiologist Assistant; EMT = Emergency Medical Technician

### Variation in App Use with Known Markers of Surgical Case Volume

Figure 1a demonstrates that there was consistent data collection from users over the study period. In the US (Figure 1b), reductions in app use occurred on major holidays, including the period around US Thanksgiving Day (i.e., the fourth Thursday of November) and Christmas Day. This is consistent with case volume data obtained from University of Washington Medical Center and Seattle Children’s Hospital (Online Supplement, Figure S2) and has been previously described in the literature as well.^25^ In Muslim-majority countries (Figure 1c), no decrease was seen on US Thanksgiving Day, and a smaller decrease was observed around Christmas Day, but there were large reductions in app use during the month of Ramadan and around the time of Hajj. In Brazil (Figure 1d), which has a large app user base, there was a notable decrease in app use during the Carnival celebration in 2019 and 2020 and the Corpus Christi celebration in 2019. There was also expected variation in app use by day of week, consistent with known data demonstrating that surgical case volumes are highest during the middle of the week and much lower on weekends (Online Supplement, Figure S3).^26^

### Impact of COVID-19 on Surgical Case Volumes

The impacts of COVID-19 on app use are also illustrated in Figure 1. Notably, all regions demonstrate steep declines in app use following the WHO declaration of global pandemic status. In Muslim-majority countries, there may have been declines in app use slightly prior to this declaration. Globally, there does appear to be an increase in app use that occurred in the last several days of data included in this sample.

We further parsed this global data to illustrate variability in app use relative to COVID-19 case counts. Overall, app use declined as COVID-19 cases mounted during the examined dates (Figure 2). Region-specific findings include:

1. In Sub-Saharan Africa (Figure 2a) and World Bank Low Income (Figure 2b) countries, the decrease in app use continues on a downward trend. The number of daily COVID-19 cases in Sub-Saharan is likely vastly underestimated due to lack of testing.
2. In the East Asia and Pacific World Bank region (Figure 2c), a decrease in app use in February was concordant with the initial rise of new cases in that region. App use rebounded and then decreased again following the WHO declaration.
3. In World Bank High Income Countries (Figure 2d; Figure 3a,b,d), the reductions in app use appears to have plateaued, with a possible recent increase in app use.
4. In Italy (Figure 3a) and Germany (Figure 3b), decreases in app use were seen well before the WHO pandemic declaration. By that time, the app use in Italy had already reached its current plateau. The decrease in app use in Germany was not nearly as pronounced as in Italy.
5. In India (Figure 3c) and the USA (Figure 3d), decreases in app use followed the WHO declaration and were abrupt. App use in India appears to be increasing; has reached a plateau in the US.

**Figure 2:**
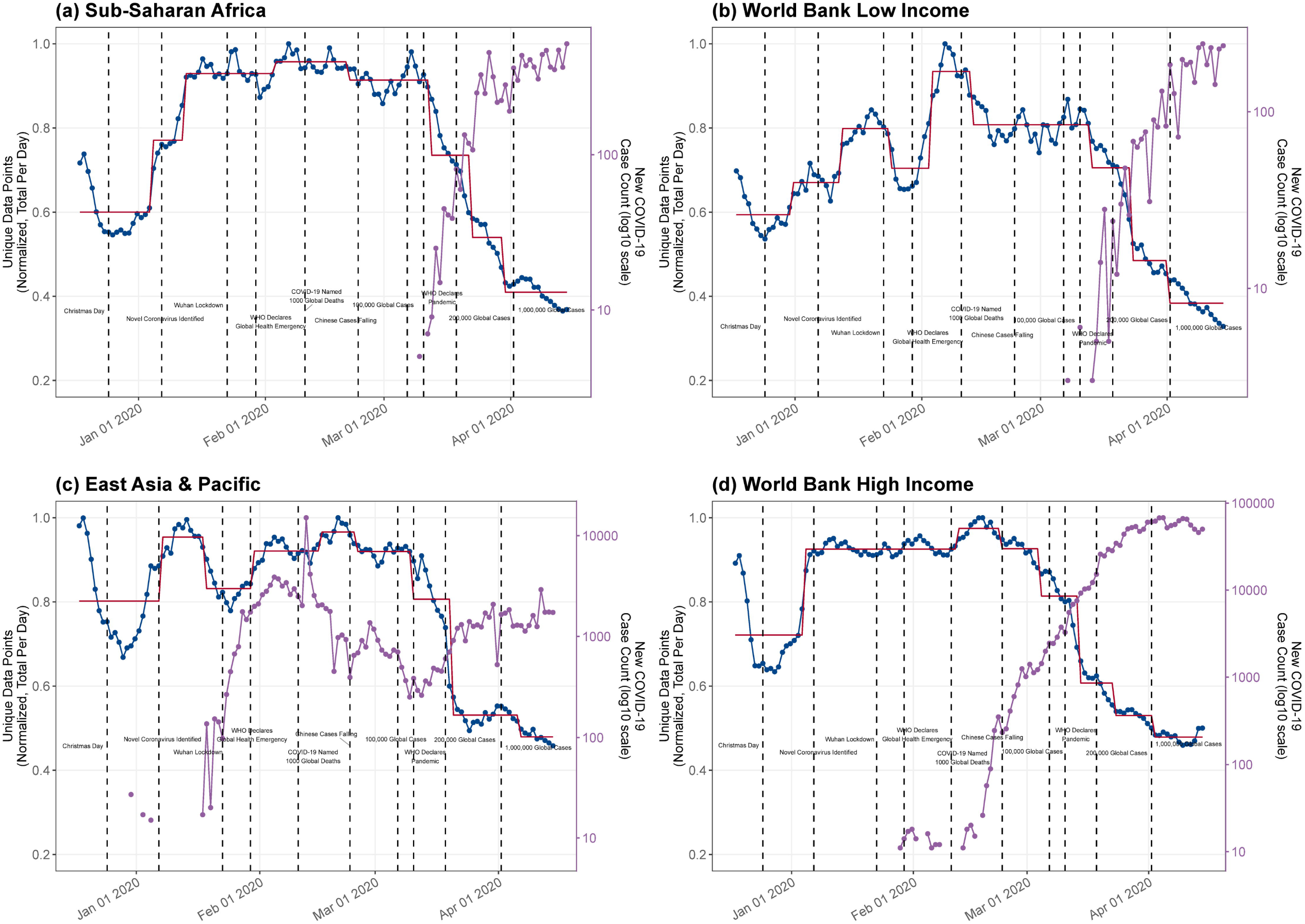
The impact of COVID-19 on app use as a proxy for surgical case volumes, plotted with daily new COVID-19 case counts, in specific groups of countries as defined by the World Bank: (a) Sub-Saharan Africa, (b) World Bank Low income countries, (c) East Asia & Pacific, (d) World Bank High income countries.

**Figure 3:**
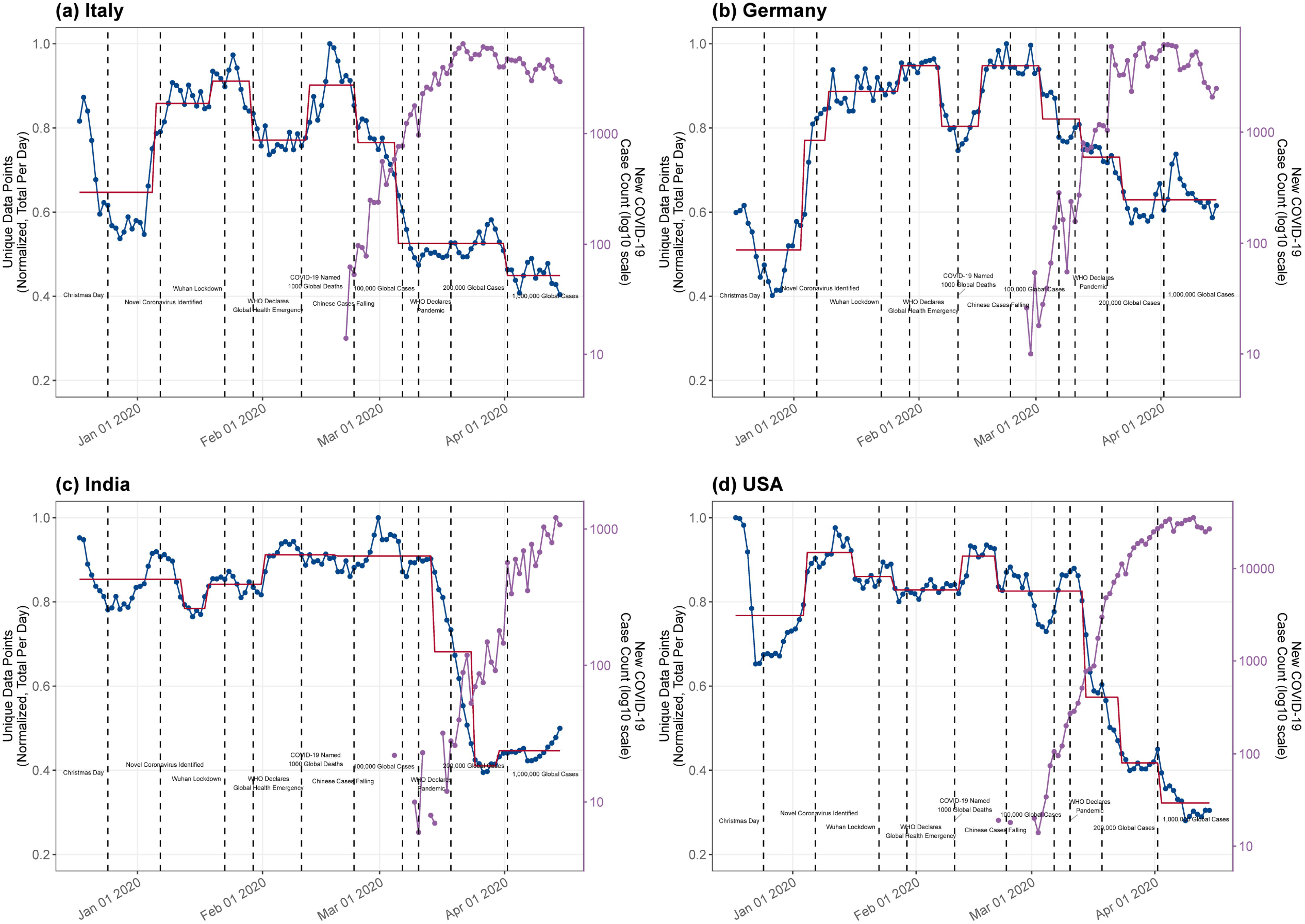
The impact of COVID-19 on app use as a proxy for surgical case volumes, plotted with daily new COVID-19 case counts, in specific countries: (a) Italy, (b) Germany, (c) India, (d) USA.

A choropleth of the global impact of COVID-19 on app use was generated (Online Supplement, Figure S4) and further demonstrates the near universal reductions in app use observed. Overall, by country there was a median global reduction in app to 58% of baseline (interquartile range, 46%-75%), with reductions as low as 11% of baseline use in some countries. There was no clear relationship between overall reduction in app use and COVID-19 case count at the end of the study period in the aggregate (Online Supplement Figure S5). When examining the cross-correlation function of COVID-19 case count versus reductions in app use in individual regions and countries, there was strong inverse correlation between these two time series at very low lags (not shown).

## Discussion

In the present work, we demonstrate that app metadata provides a low-cost, novel approach to the qualitative assessment of surgical case volume, particularly in countries where such real-time data may be difficult or impossible to otherwise obtain. The substantial reductions in app use observed in relation to major holidays and day of week correlate well with known impacts on surgical case volumes. ^25–28^ This methodology similarly demonstrated previously unreported but plausbile reductions in surgical case volumes during Ramadan and Hajj in Muslim-majority countries and during Corpus Christi and Carnival in Brazil. As a proxy for surgical case volumes, this app data demonstrates expected but troubling impacts of the COVID-19 pandemic worldwide.

Globally, surgical case volumes are difficult to assess.^7,8^ The difficulties in obtaining this and related data have been identified as a major challenge by the Lancet Commission on Global Surgery in the seminal Global Surgery 2030 report.^6^ In a recent modeling study of global surgical volumes, less than a third of countries could offer data to support the analysis.^7^ Even where such data are available from public health ministries, it is not available in real-time and may be limited to healthcare delivered by governmentally funded facilities.^29^

The benefits and challenges associated with collection of healthcare data using mobile technology have been previously discussed.^30,31^ Benefits include a decentralized approach to collection of data from the 95% of the global population living in an area covered and subscribed to mobile cellular service.^32^ Specifically in LMIC, there were 5.5 billion mobile phone subscriptions, representing nearly 92 subscriptions per 100 inhabitants.^33^ Challenges include dissemination of specific applications, the types of data that can be collected, the tradeoff between apps that have clinical utility and the data that can be gleaned from the use of those apps, the availability of multiple platforms (e.g. Android, iOS), and the analysis and interpretation of stochastic app use data.

The impact of the COVID-19 pandemic on surgical capacity in LMIC may be dire. A large number of people are without adequate access to safe anesthetic and surgical care at baseline.^6^ There is the potential that this pandemic will further deplete resources of already-stressed LMIC healthcare systems, worsening access to surgical care. Our data indicate that the impacts of COVID-19 on surgical volumes have been substantial; it will be of great value for governments, global health organizations, and philanthropic organizations to have access to data providing markers of recovery of surgical capacity -- or lack thereof -- in LMIC. Global strategies could be put in place by large organizations (such as the WHO) to attempt mitigation. The lack of association between COVID-19 case counts and reductions in app use are likely explained by the fact that hospital and governmental policies are likely driving surgical scheduling behavior to a greater extent than the actual patient surge in most places.

Practical utilization of the app from which data were gathered and analyzed is a specific strength of the present work. Users download and use the app for the clinical care of patients. The app was never advertised, nor was its use encouraged, through notifications or other mechanisms. Thus, app use reflects, on an individual basis, stochastic clinical care events. This same stochasticity highlights a limitation of the work, namely that data from individual regions or countries with a small user base reduces the confidence we can assign to the association between app use time series data and surgical case volume. Fortunately for global health purposes, our previous work demonstrates that, in LMIC, the app has greater frequency of use as compared to HIC.^27^

Another important limitation of the present work is the qualitative nature of the interpretation that can be made of shifts in app use. The app is used primarily for the care of pediatric cases (~75% of app uses are for patients age 12 and younger), and users also consult the app during emergencies.^34^ Thus, the pattern of app use is greater on weekends (when a greater proportion of cases are emergencies) than would be expected based on the actual proportion of surgical case volumes comparing weekdays to weekends. This pediatric clinical predominance may drive greater use of the app in LMIC where (a) subspecialty training in pediatric anesthesia is less prevalent compared to HIC and (b) as high as 50% of the population may be under the age of 16. Patterns in app use will additionally be biased towards impacts specifically on pediatric case volumes. Notably, these needs in LMIC are not trivial: 1.7 billion children lack adequate access to surgical care and an estimated 85% of children in LMIC will need surgical care by age 16.^35^ Conversely, app utilization patterns may be relatively less impacted in HIC with dedicated pediatric hospitals. Given the differential impact of SARS-CoV-2 in the young versus the aged, and the proportion of elective versus non-elective surgery in pediatric patients, a greater degree of pediatric surgery may be seen compared to surgery for adults.

In conclusion, we present a real-time qualitative monitor of the impact of COVID-19 on global surgical volumes, particularly in at-risk LMICs. There is currently a clear trend of decreased surgical volume in certain regions associated with the onset of the COVID-19 pandemic. The trends in different regions will likely change with time depending on pandemic surge timing, restoration of normal operations, baseline healthcare capacity, and resource allocation. Combined with other information sources, this tool provides the opportunity to monitor and act in real time on data to identify areas continuing to suffer from depressed surgical volumes and direct resources to areas with the most need. A strategic global approach will decrease the morbidity and disability associated with diminished surgical capacity. Longer term, this tool can be combined with other data to assist with measurement of global surgical capacity as part of the Global Surgery 2030 initiative. As such, a near real-time dashboard has been developed (http://globalcases.info) and deployed as part of these efforts to ensure ongoing accessibility of this information.

## Data Availability

Data used and code written for the present analysis shall be made upon reasonable request for academic purposes.

http://globalcases.info

## Sources of support

NIH/NIGMS (T32 GM086270-11, DRL

I, Vikas O’Reilly-Shah, had full access to all the data in the study and had final responsibility to submit for publication.

## Author contributions

All authors meet ICMJE criteria for authorship.

V-OS conceived the work, designed data collection tools, monitored data collection, wrote the statistical analysis plan, cleaned and analysed the data, and drafted and revised the manuscript. He is guarantor.

VM interpreted the data, and drafted and revised the manuscript.

FME interpreted the data and revised the manuscript.

JES interpreted the data and revised the manuscript.

WVC interpreted the data, and revised the manuscript.

DRL analyzed the data, interpreted the data, and revised the manuscript.

NJK interpreted the data and revised the manuscript.

EH interpreted the data and revised the manuscript.

CSJ interpreted the data and revised the manuscript.

## Acknowledgements

The authors would like to thank Alexandra Torborg, MBChB (University of KwaZulu-Natal, Nelson R Mandela School of Medicine) for insightful thoughts and comments on the work and the manuscript.

